# Pregnancy outcomes and long-term patient and graft survival after kidney transplantation

**DOI:** 10.1101/2022.01.18.22269382

**Authors:** Eloísa Radaelli, Gisele Meinerz, Lázaro Pereira Jacobina, Rosana Mussoi Bruno, Valter Duro Garcia, Eduarda Tanus Stefani, Letícia Kortz Motta Lima, Natália Klaus Weber, Elizete Keitel

## Abstract

**Introduction:** Kidney transplant is considered the method of choice in the treatment of end-stage chronic kidney disease, improving survival and quality of life, and often allowing the recovery of fertility among woman. Objective: This study aims to assess the outcomes of post-transplant pregnancy, newborn and patient graft survival, compared to a control group of transplanted women who weren’t pregnant.

**Methods:** A retrospective case-control study was conducted at the Kidney Transplant Service of Santa Casa de Misericórdia in Porto Alegre, among 1.253 female patients who underwent kidney transplantation and were of childbearing age between 1977 and 2016. They were compared to a control group matched for age, type of donor, date of transplant and immunological risk.

**Results:** 76 (6,1%) of the patients became pregnant, resulting in a total of 93 pregnancies. Compared to the control group, at 10 years after transplantation there was no significant difference in relation to graft loss (74% vs 66%, p=0.524), and the survival of pregnant patients was higher (97% vs 79%, p=0.018). We found that 40% pregnancies progressed to abortion, 30% of the deliveries were preterm, and 26% happened at term. Three pregnancies progressed to stillbirth. Of the 56 pregnancies that evolved, 23 (41%) resulted in pre-eclampsia, and one in eclampsia.

**Conclusion:** 6,1% of the women in the study became pregnant, and 56% of these pregnancies were successful. Compared to the control group, we found that those who got pregnant had similar graft survival outcomes and longer patient survival.

## INTRODUCTION

Kidney transplant is considered the treatment of choice for end-stage chronic kidney disease. Brazil is the second country in number of kidney transplants, and many hospitals and specialised services are certifiably capable of performing the procedure in Brazil. Among them is Santa Casa de Misericórdia in Porto Alegre, with over 5.000 thousand kidney transplants up to 2019. Over 2.000 of these patients were women, many of which were of childbearing age.

In addition to the general improvement in quality of life, women often regain fertility and shortly find it possible to become pregnant^1^. With the advances in surgical techniques and immunosuppression therapy, there has been an increase in successful pregnancies among these patients^2^. The first successful pregnancy in a kidney receiver was described in 1958 and, since then, many other were reported. Nevertheless, the incidence of non-complicated pregnancies is still much lower than in other women.

Pregnancy viability and maternal complications are still an important subject of study. Some data show that, among female kidney recipients, 35% of gestations do not progress beyond the first trimester, due to spontaneous miscarriages or therapeutical abortions; after the first trimester, the rate of success is higher. Risk factors for the development of graft rejection in these patients include high levels of previous serum creatinine and fluctuating levels of immunosuppressive drugs, caused by pregnancy-related changes in the distribution and clearance of these agents^3^. Potentially significant complications include hypertension, pre-eclampsia, rejection and infection. For the foetus, the risk of prematurity is the main factor to be considered, as well as low birth weight and intrauterine growth restriction ^4^. The most important prognostic factor for favourable outcomes in these patients is good previous renal function and the absence and/or adequate control of hypertension^5^. Several studies in literature address this topic, describing the adverse effects of pregnancy for both the woman and the fetus; however, few of them compare the outcomes of mortality and graft survival between pregnant women to those of women who did not get pregnant.

## METHODS

This is a retrospective case-control study developed at the Kidney Transplant Service of Santa Casa de Misericórdia in Porto Alegre, South of Brazil. Female patients of childbearing age who underwent transplantation from 1977 to 2016 and followed up until the end of 2020 were evaluated. Patients not followed up for at least one month after the transplant were excluded. Pregnant patients were compared with non-pregnant controls, being matched for age, donor type, date of transplant and immunological risk. The variables collected for analysis of pregnant women included the time between transplantation and pregnancy, renal function, presence of proteinuria above 500mg/day, systemic arterial hypertension, immunosuppressive regimen, and pregnancy outcome (spontaneous abortion, therapeutic abortion, type of delivery and delivery time). In the newborn, birth weight, gestational age and evolution after birth (stillbirth) were evaluated. After pregnancy, the frequency of episodes of acute rejection and renal function were assessed. Patient and graft survival were both compared with the control group.

Based on a previous study, the sample power was calculated assuming that there may be a 10% graft survival difference in 5 years after transplantation between pregnant and non-pregnant patients. We have a power of 77.3%, with a significance level of 5% (two-tailed) to detect this difference, performing a case-control study in a 1:1 ratio with a group of 65 patients in each group. The calculation was performed using the WinPepi program, version 3.85 (Power of test for difference between proportions – module P1). The study was approved by the Hospital’s Research Ethics Committee. Data from medical records were analysed using the Statistical Package for Social Sciences (SPSS) version 21.0 (SPSS INC., Chicago, IL). Categorical variables were analysed using the chi-square test and continuous variables using Student’s t test. Patient and graft survival were assessed using the Kaplan-Meier method. Data normality was verified using the Kolmogorov-Smirnov test with Lilliefors test correction. Continuous variables were presented as median and interquartile range or mean and standard deviation, categorical variables as absolute and relative frequencies. Comparisons between groups were performed using the Mann-Whitney U test. Intragroup comparisons were performed using the Wilcoxon paired test. A significance level of p < 0.05 was considered for all analyses.

## RESULTS

During the study period, 1.253 women were of reproductive age and had a minimum follow-up of one month. Forty (11.7%) patients became pregnant among the 341 transplanted from 1977 to 1999, and 36 (3.94%) among the 912 transplanted from 2000 to 2016. A total of 76 (6.1%) were pregnant; 65 of them had a single pregnancy, 6 had 2 pregnancies, 4 had 3 pregnancies and 1 had 4 pregnancies during the study period, resulting in a total of 93 pregnancies analysed. In the control group, 76 patients matched for age, type of donor, date of transplantation and immunological risk were included.

Table 1 shows the demographic data of the groups. Table 2 offers a description of the pregnant patients. We observed a median time between transplantation and conception of 48 months. The creatinine value of pregnant women before conception and up to one year after delivery did not change significantly (mean creatinine was 1.2mg/dL) and proteinuria before pregnancy was an average of 0.3g/dL. Sixty-seven percent of the pregnant women had hypertension and 47% of them had more than one risk factor for poor outcomes associated with pregnancy (systemic arterial hypertension, serum creatinine >1.5mg/dL, proteinuria, gestation time less than 2 years after transplantation). Twenty-three (41.8%) presented preeclampsia and one of them had eclampsia.

**Table 1:** Demografic charateristics of cases and controls

| Variable | Pregnant n =76 | Control n=76 |
| --- | --- | --- |
| Mean age at transplant (SD) | 23.81(7.29) | 25.45 (6.01) |
| Live donor (%) | 52 (68) | 52 (68) |
| Fist Transplant | 68 | 67 |
| Diabetes | 1 | 1 |
| Donor specific antibodies | 4 | 3 |
| Induction therapy (depleting) | 7 | 6 |
| Maintenance Immunosuppression |  |  |
| Azathioprine | 51 | 51 |
| Mychophenolate acid | 16 | 22 |
| Cyclosporine | 39 | 41 |
| Tacrolimus | 16 | 19 |
| Steroids | 72 | 74 |
| imTOR | 1 | 0 |
| 1977-1999 | 40 | 42 |
| 2000- 2016 | 36 | 34 |

**Table 2:**
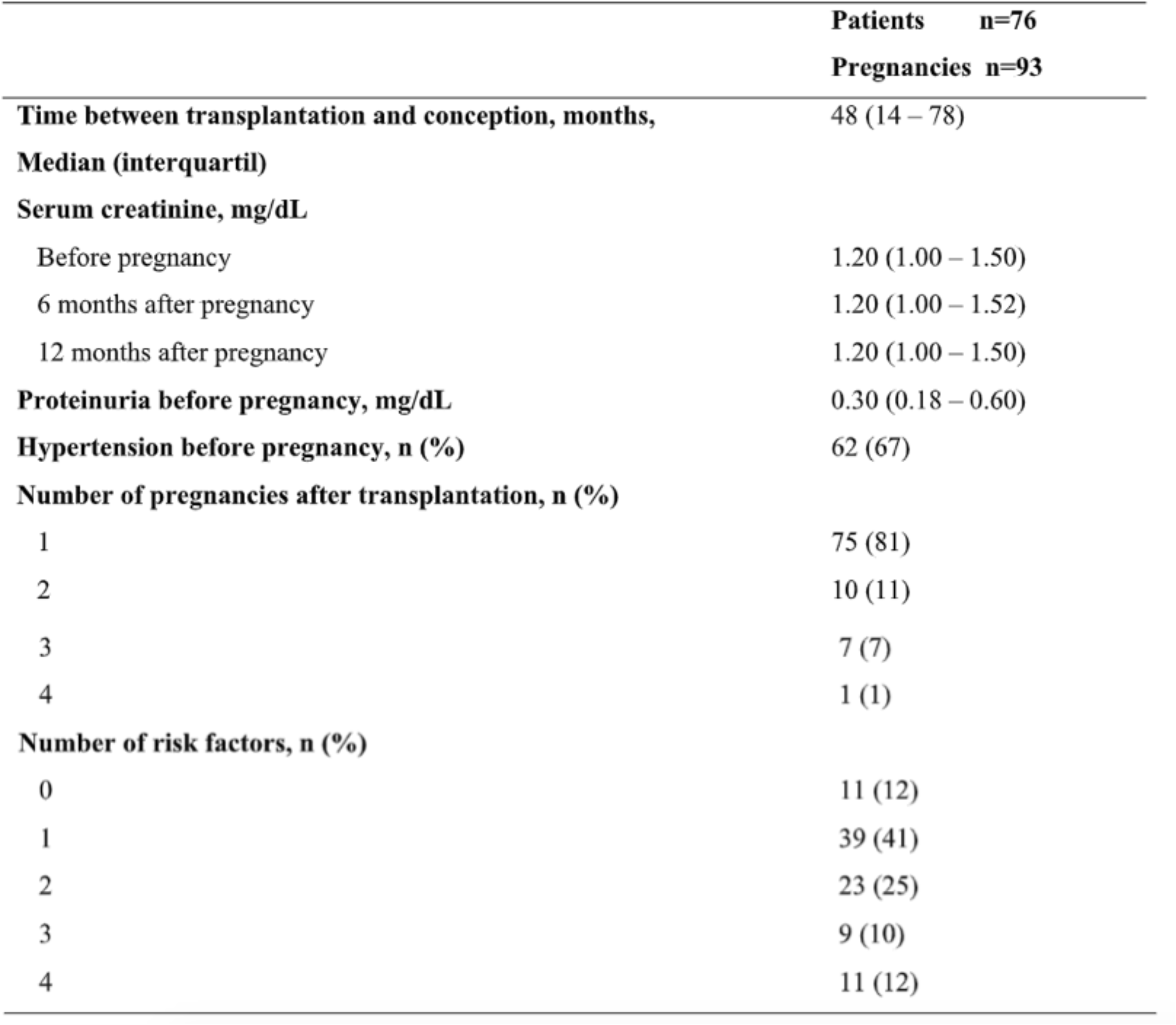
Pregnancy data

Regarding pregnancy outcomes, as described in Table 3, we observed that a total of 40% of pregnancies progressed to abortion (both spontaneous and therapeutic). Most therapeutic abortions (n=16) occurred before the year 2000. Pregnancy failure (n=41) was associated with the presence of two or more risk factors (OR: 4.67 CI 1.69 – 10.76 p=0,001) and graft loss within 2 years after pregnancy (n=13) was associated the presence of three or more risk factors (OR: 5.83 CI 1.68-20.21 p=0.003). The rate of graft rejection observed among the 76 pregnant women was 15%. In 30% of pregnancies the births were preterm and 26% were at term. Three pregnancies progressed to stillbirth. Only 5 (9.1%) had vaginal delivery. The average weight of live births was 2.400 grams.

**Table 3.** Pregnancy and graft outcomes

|  | Pregnancy (n=93) |
| --- | --- |
| **Pregnancy outcome n (%)** |  |
| Live births | 52 (56) |
| Miscarriage | 19 (20) |
| Therapeutic terminations | 19 (20) |
| Stillbirth | 3 (4) |
| **Live birth** |  |
| Preterm births | 28 (30) |
| Born term delivery | 24 (26) |
| **Preeclampsia/eclampsia n (%)** | 24/ 55 (43.63) |
| **Birth weight of infant (g)** | 2400 (1925 – 2750) |
| **Post-pregnancy graft rejection, n (%)** | 11 (15)<sup>*</sup> |
| **2-years post-pregnancy graft loss, n (%)** | 13 (14) |

Values are presented as median (range) unless otherwise noted.
<sup>\*</sup>n=76

Pregnant women compared to the control group, at 10 years after transplantation, had no significant difference related to graft loss (74% vs 66%, p=0.524), but the survival of pregnant patients was higher (97% vs 79%, p= 0.018). The results of the comparison of patient and graft survival between pregnant women and the control group, including patients who died with a functioning graft, are shown in Graphs 1, 2 and 3.

**Graphic 1:**
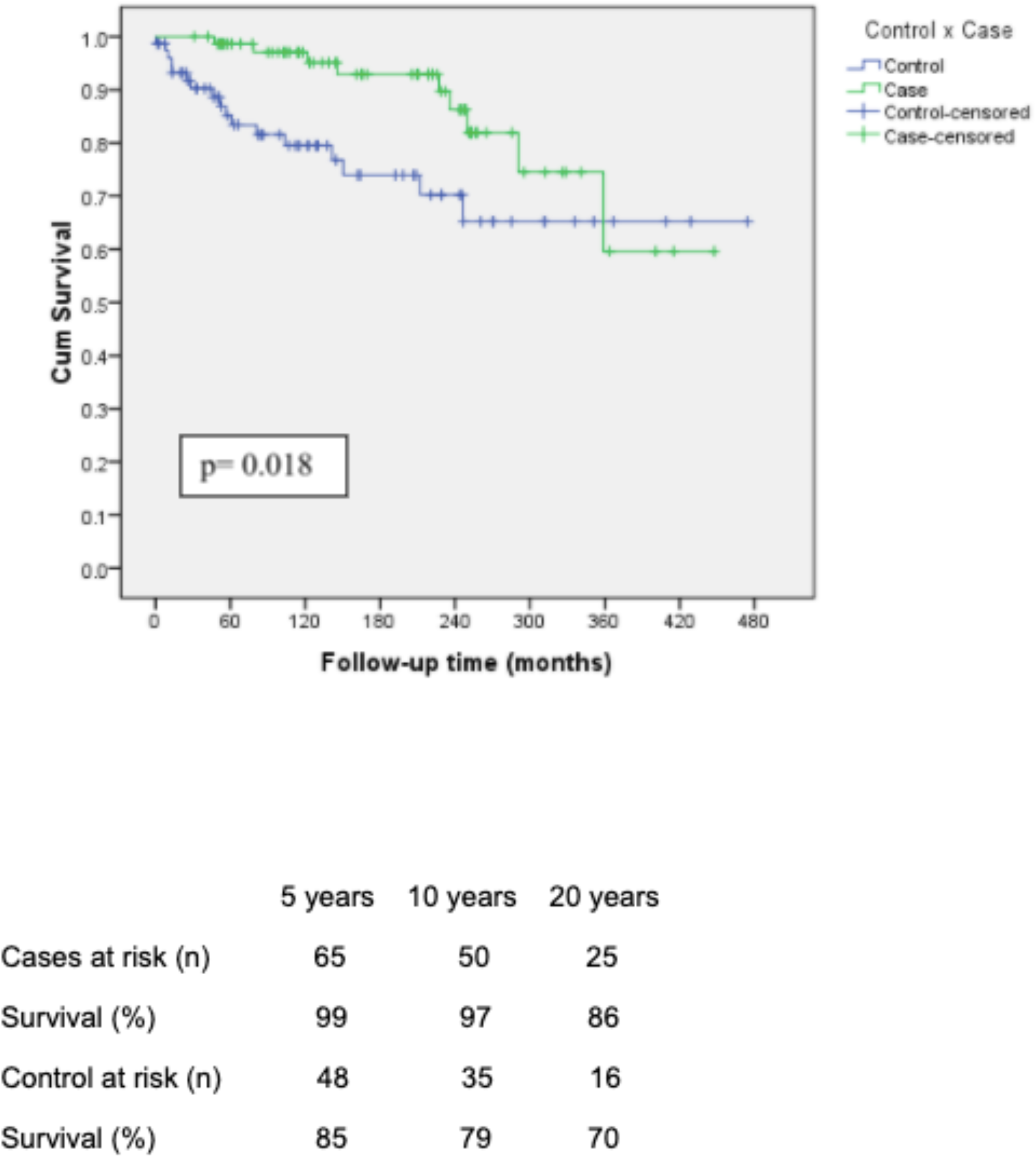
Patient survival comparing pregnant patient with control

**Graphic 2:**
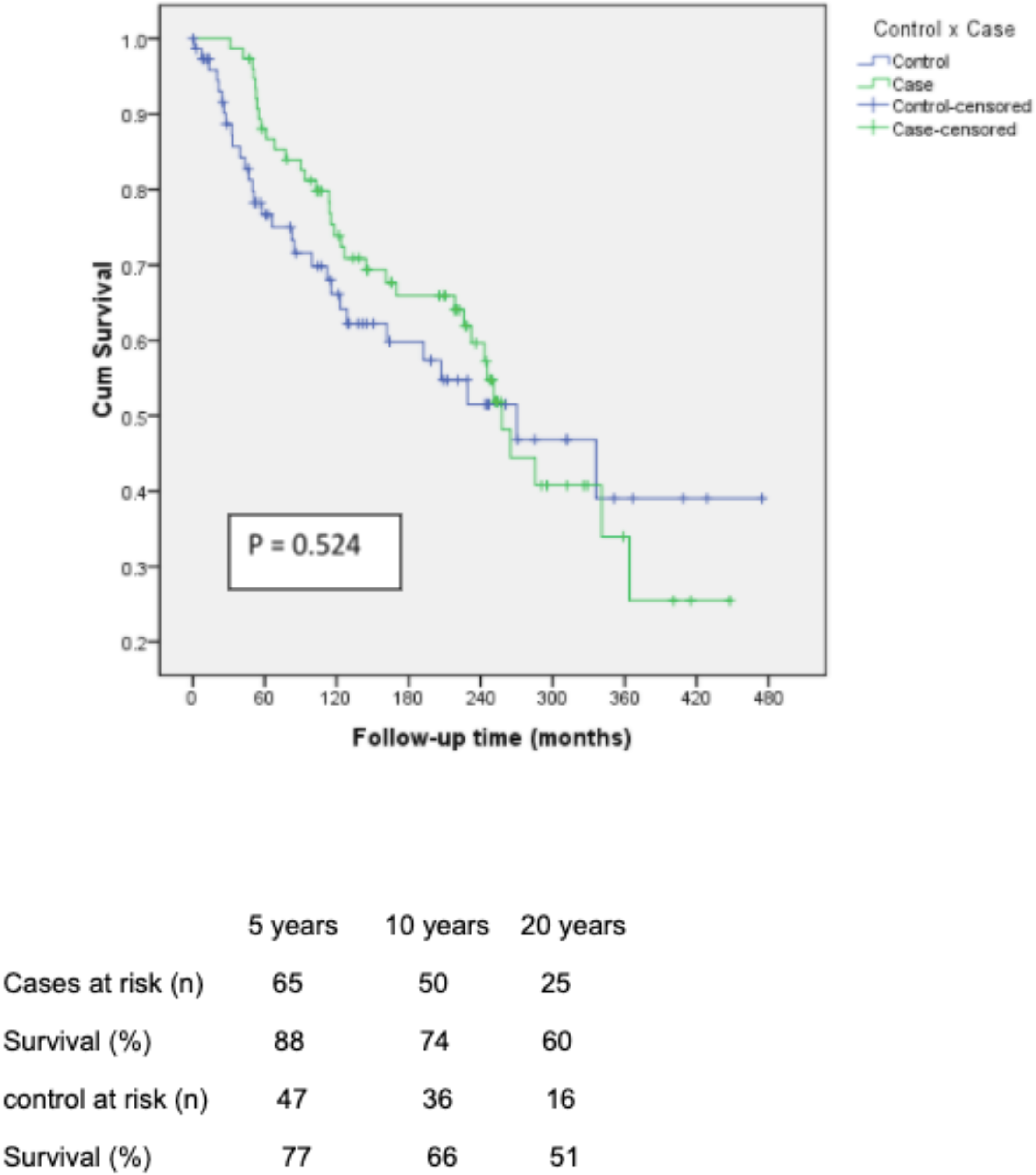
Death censored graft survival comparing pregnant patient with control

**Graphic 3:**
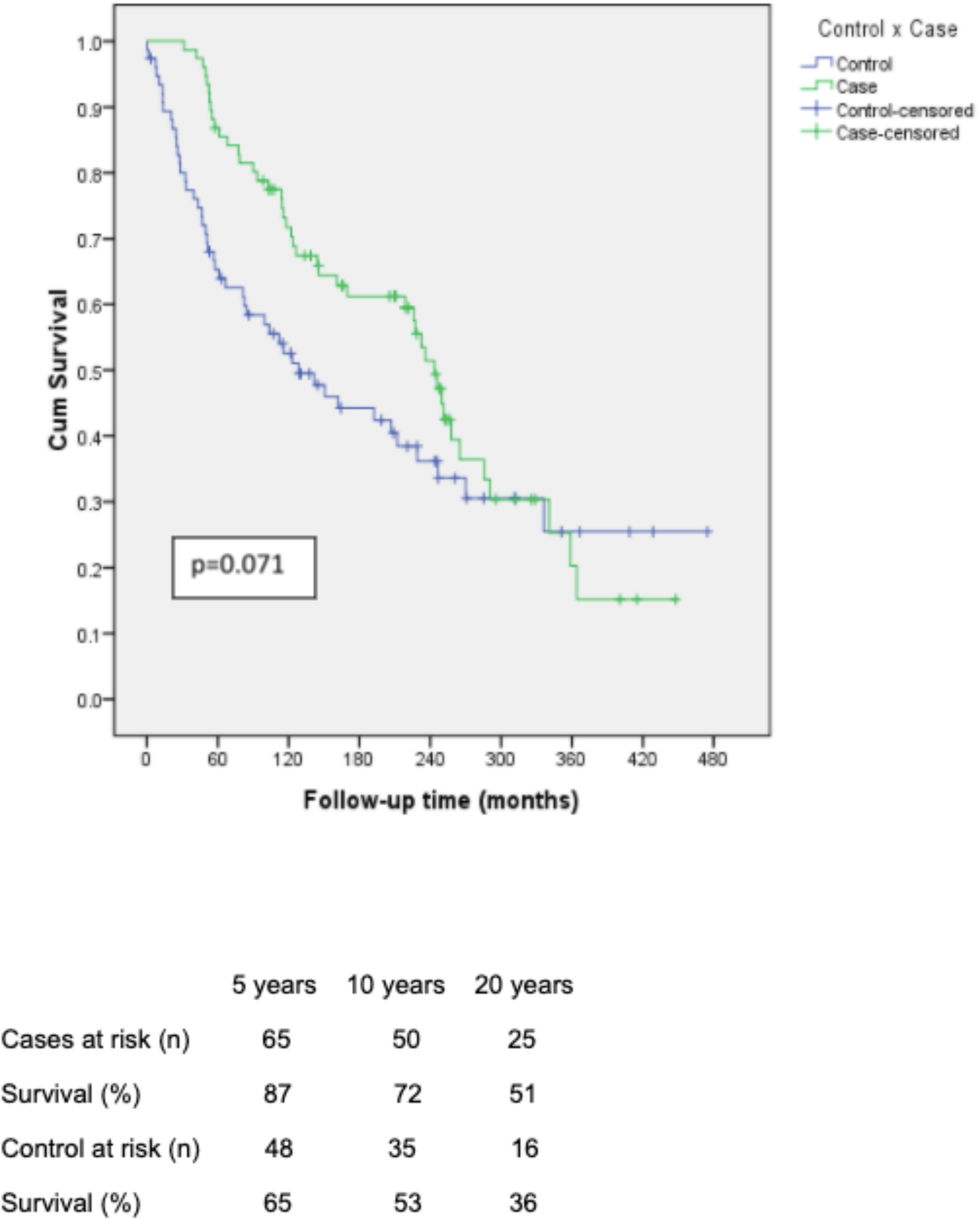
Graft survival non death censored comparing pregnant patient with control

## Discussion

Fertility recovery is a demonstration of improved overall health after kidney transplantation. However, pregnancy after kidney transplantation is still considered a high risk for both mother and child. In our study, we observed that the frequency of pregnancy decreased over the years, which is probably due to a multidisciplinary counseling program established at the Transplantation Service. In our sample of 93 pregnancies in 76 women, only 12% had no risk factors prior to pregnancy. The median time was 48 months after kidney transplantation, in line with what is recommended in the literature, which advises a period of 24 months between transplantation and pregnancy, not exceeding five years due to the risk of deterioration of graft function^6^. It is known that the most important prognostic factor for favourable pregnancy outcomes in transplant patients is a good previous renal function and the absence and/or good control of hypertension, maintaining serum creatinine < 1.5 mg/dL with no or minimal proteinuria ^7^. Regarding these variables, we observed that the mean pre-conception creatinine at six and twelve months after delivery remained stable. On the other hand, systemic arterial hypertension was the most prevalent comorbidity found in our pregnant patients (67%). According to the literature, the prevalence of hypertension in pregnant women with chronic kidney disease ranges from 27% to 40% and it is an independent risk factor for adverse pregnancy outcomes. Women with chronic kidney disease have ten times the risk of pre-eclampsia when compared to women without kidney disease. In post-transplant women, pre-eclampsia occurs in up to 40% of pregnancies^8^, similarly to our findings. The relationship between pre-eclampsia and chronic kidney disease is characterised by a set of manifestations that include hypertension, proteinuria, impaired renal function and increased cardiovascular risk. It is mportant to note that in the only case of eclampsia in our sample, the patient did not have any risk factor prior to conception and eventually died as a consequence of it. Our policy is to share the patient care with a high-risk obstetrician team.

Among the concerns of pregnancy after kidney transplantation is the risk of graft loss. The presence of 3 or more pre-pregnancy risk factors was associated with graft loss within 2 years of pregnancy. Graft dysfunction, defined as an increase in creatinine of 0.3 mg/dL or more, can occur during pregnancy, but appears to be temporary in most cases^9^. In the past, it was believed that immunological changes during pregnancy contributed to a lower incidence of rejections during this period. Recent literature reports that the incidence does not differ in relation to the general population of transplant recipients, being reported in about 4-5% ^10^. In our sample, we observed a frequency of 11%. According to our results, we observed favourable pregnancy and graft outcomes especially in pregnant women with absence of risk factors, or presence of only one risk factor. Thus, pre-gestational counselling is extremely important since modifications of the immunosuppressive regimen are often necessary before conception^6^, as well as risk assessment. However, we observed a similar long term graft survival to the paired control group and even more a higher rate of patient survival. Similar findings has been reported in literature.

Higher cesarean section rates have been reported and our results are in agreement with it. In addition, the need for blood transfusion, anemia, ectopic pregnancy, infections and gestational diabetes^11^ are reported, but were not evaluated in our study. Regarding fetal complications, the literature presents as main events preterm birth, restricted intrauterine growth, low birth weight, acute respiratory distress syndrome, infections, adrenal insufficiency, chromosomal abnormalities, thymus atrophy, leukopenia, thrombocytopenia, anemia, hydronephrosis and malformations^12^. Abortion is present in up to 26% of pregnancies and may be associated with high rate of maternal risk factors and the use of immunosuppressive medication^13^. Our sample had a higher rate of therapeutic abortion prior to the year 2.000, probably due to the greater number of unplanned pregnancies in patients with many risk factors. There was a significant therapeutic abortion reduction with the improvement of multidisciplinary counselling and an adjustment program of immunosuppression prior to conception^14^. The overall high rate of abortion lead to a lower rate (56%) of successful deliveries, compared to the 75% rate currently published^15^. Although our sample included patients with a wide range of transplant era, the pregnancies occurred after 2.000 also had a similar rate of success due to increase in spontaneous abortion. According to a recent meta-analysis published in 2020^16^, graft survival is similar between the groups of pregnant and non-pregnant women, which corroborates our findings, however, survival among patients was not evaluated in the study mentioned.

We observed that prematurity is the main unfavourable perinatal outcome. It may lead to several consequences in the future, being the main cause of perinatal mortality, and it results from several factors, including the premature rupture of the amniotic membrane, the chronic use of corticosteroids, the high incidence of urinary tract infections and obstetric interventions^14^. Of our 93 pregnancies analysed, 30% were premature and 26% of them were at term, which is in line with data from pre-existing studies. This happens because delivery is usually induced before 37 weeks due to maternal complications. Regarding fetal loss, three pregnancies resulted in stillbirth. Some risks has been directly associated to it, such as afro-descendancy, diabetes as a cause of end-stage renal disease and low family income^14^.

## Data Availability

All data produced in the present study are available upon reasonable request to the authors

## Notes

### Competing Interest Statement

The authors have declared no competing interest.

### Funding Statement

This study did not receive any funding

### Author Declarations

Ethics committee of Irmandade da Santa Casa de Misericordia de Porto Alegre - ISCMPA gave ethical approval for this work

